# A multicenter prospective validation cohort does not confirm the diagnostic yield of [^18^F]FDG PET/CT imaging in kidney allograft subclinical rejection

**DOI:** 10.64898/2026.03.07.26347838

**Authors:** Pierre Lovinfosse, Antoine Bouquegneau, Annick Massart, Lissa Pipeleers, Catherine Bonvoisin, Laurens Carp, Hendrik Everaert, Alexandre Jadoul, Amélie Dendooven, Caroline Geers, Stephanie Grosch, Pauline Erpicum, Rachel Hellemans, Laurence Seidel, Laurent Weekers, Roland Hustinx, François Jouret

## Abstract

**Background:** Subclinical kidney allograft acute rejection (SCR) corresponds to “the unexpected histological evidence of acute rejection in a stable patient”. The diagnosis of SCR relies on surveillance biopsy. Positron emission tomography (PET/CT) after injection of F^18^-fluorodeoxyglucose ([^18^F]FDG) has been proposed as a non-invasive screening approach. In the present multicenter prospective study, we assess the diagnostic yield [^18^F]FDGPET/CT to rule out SCR in stable KTR at 3 months *post* KTx.

**Methods:** From 01/2021 to 03/2025, we prospectively combined surveillance biopsy and [^18^F]FDGPET/CT at ∼3 months *post* transplantation in adult kidney transplant recipients from 4 independent imaging centers. The mean standardized uptake value (mSUV) was measured in kidney cortex and referenced as a ratio to psoas muscle mSUV (mSUVR).

**Results:** Our multicentric 185-patient cohort was categorized upon Banff-2022: normal (n=158); borderline (n=18); SCR (n=9, including 6 T-cell-mediated rejection and 3 microvascular inflammation). No significant correlation was observed between the mSUVR and ti score (R=0.032, p-value=0.67). The mSUVR reached 2.33 [1.97-2.93], 2.71 [2.50-3.33] and 2.42 [2.27-3.14] in normal, borderline and SCR groups, respectively. In multivariate models stratified by center, the risk of non-normal histology (n=27, including borderline and SCR) increased with donor age (OR=1.05 [1.01-1.1], p=0.02) but not with the mSUVR (OR=4.11 [0.91-18.48], p=0.07). The risk of biopsy-proven SCR (n=9) was not significantly associated with mSUVR.

**Conclusions:** The mSUVR of [^18^F]FDG PET/CT does not reliably rule out SCR on surveillance biopsy.

## Introduction

The systematic follow-up of kidney transplant recipients (KTRs) is essential to early detect transplant rejection and appropriately personnalize the immunosuppressive regimen^1^. The diagnostic gold-standard for allograft rejection relies on the Banff classification, which provides standardized histopathological criteria for the grading of allograft rejection, including the assessment of microvascular inflammation (MVI)^2^.

Subclinical rejection (SCR) has been defined as ‘the documentation by light histology of unexpected evidence of allograft rejection in a stable patient’^3^. Surveillance transplant biopsies are, by definition, required for the diagnosis of SCR^4^. Although an ultrasound-guided core needle biopsy of kidney allograft is regarded as relatively safe, it remains an invasive procedure with a ∼7% rate of complications^5^. Furthermore, inter-observer variability and sampling errors limit its benefits. In order to optimize the currently indiscriminate use of allograft biopsies in stable KTRs, non-invasive approaches could be developed as “rule out” tests, with the highest negative predictive value^6–8^. Various non-invasive modalities for the diagnosis of SCR are currently under investigation, including imaging, gene expression profiling and omics analyses of blood and urine samples^9–11^. We and others have suggested that ^18^F-Fluorodeoxyglucose Positron Emission Tomography coupled with Computed Tomography ([^18^F]FDGPET/CT) may help to non-invasively distinguish the absence of biopsy-proven acute rejction^12–15^. Hypothetically, the immunological reaction against donor antigens induces the recruitment of mononuclear leukocytes into the renal transplant, which basically corresponds to the core of the Banff classification. The boosted metabolism of these inflammatory cells could be assessed by PET quantification of the renal uptake of [^18^F]FDG. A renal mean standardized uptake value (mSUV) below a screening threshold would therefore suggest the absence of active inflammation in the allograft, thereby precluding the surveillance biopsy. In order to mitigate the impact of sensitivity disparities of various [^18^F]FDGPET/CT devices, “target-to-background” ratios can be used. On the basis of a pilot prospective monocentric 92-KTR cohort, we postulated that a ratio of kidney/psoas mSUVR lower than 2.4 would non-invasively exclude SCR, with a negative predictive value of 98%^16^. In the present multicenter validation study, we prospectively assess the diagnostic yield of this ≤2.4 mSUVR threshold to rule out SCR in stable KTR at 3 months *post* KTx.

### Patients and Methods

#### Patients

From January, 1 2021 to March, 31 2025, we prospectively performed [^18^F]FDGPET/CTimaging in stable adult KTR who underwent surveillance transplant biopsy at ∼3 months *post*KTx in 3 independent transplant centers: Universitair Ziekenhuis Antwerpen (UZA), Universitair Ziekenhuis Brussels (UZB), and Centre Hospitalier Universitaire de Liège (CHULiège). At CHULiège, [18F]FDGPET/CT images were acquired on 2 different machines (*see infra*). Patients with a probable polyomavirus nephropathy (BK polyomavirus vriemia > 3 log_10_ copies) and patients who underwent an ABO incompatible KTx were excluded. This study has been approved by the Institutional Review Board (IRB) of CHULiège under the number B707202042977, and endorsed by the IRB of UZA and UZB. Written informed consent was obtained. Kidney transplantations were performed in accordance with the Declaration of Istanbul. Our database has been registered as NCT03764124. Clinical, biological, histological and imaging parameters were systematically collected (**Table 1**).

**Table 1.**
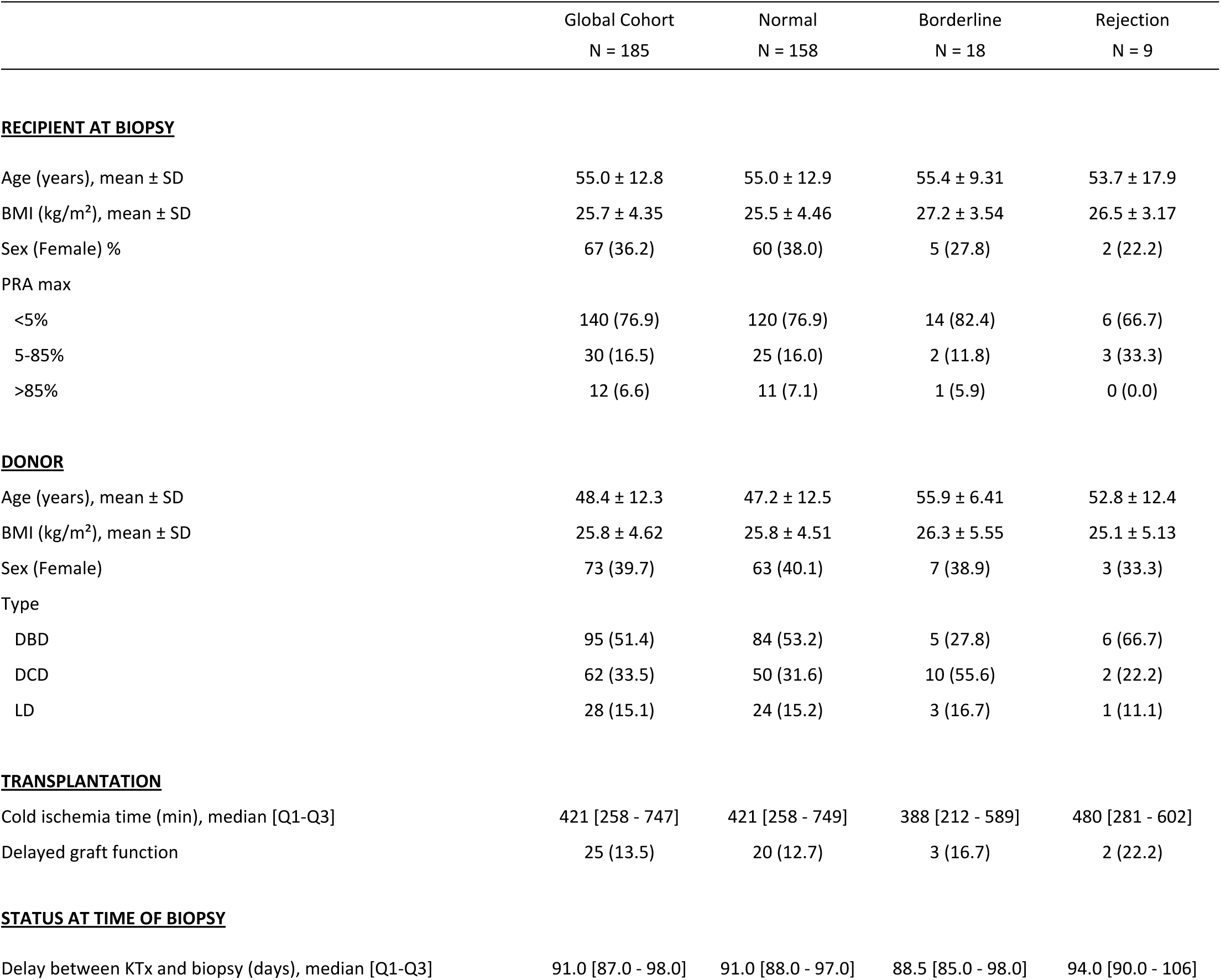
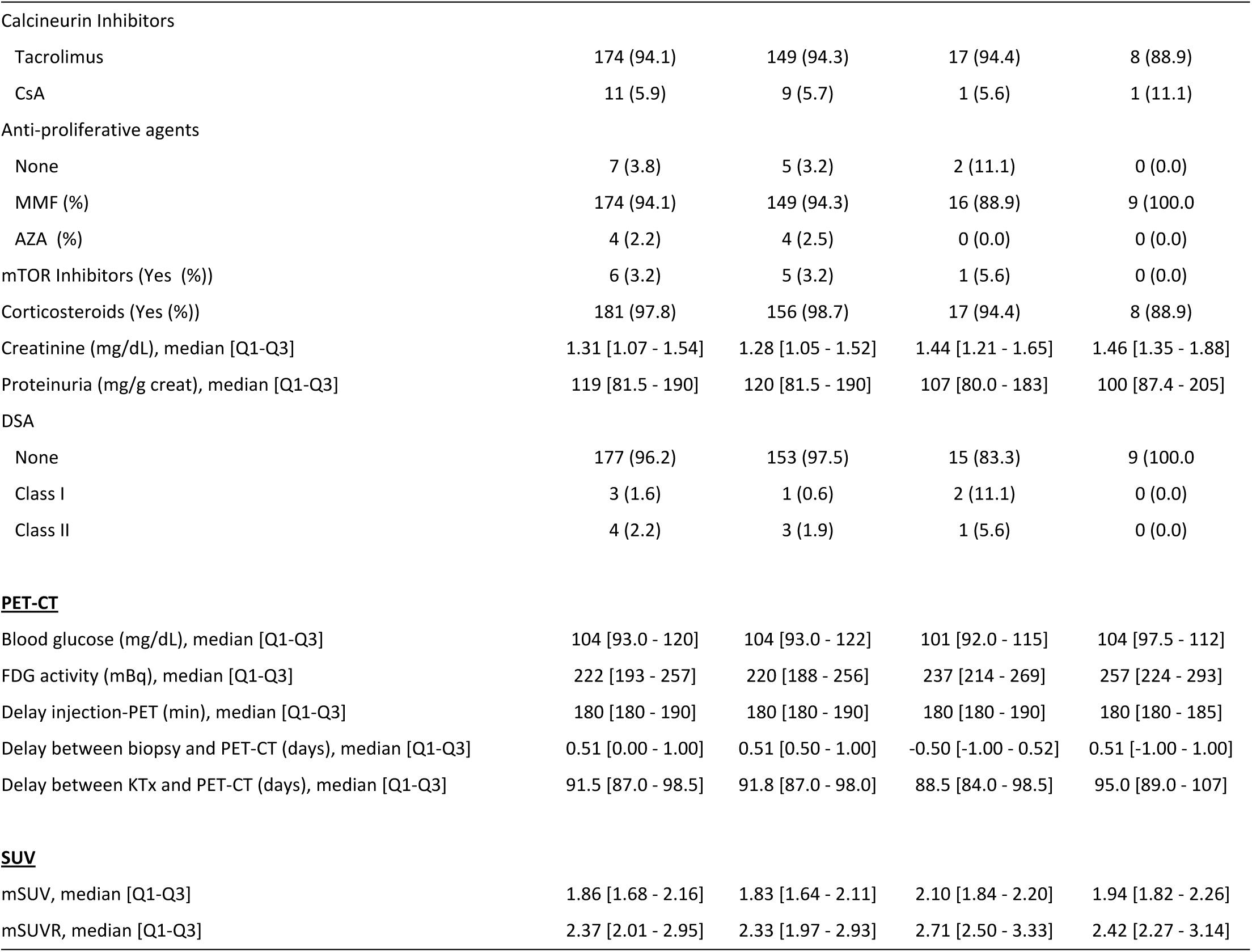
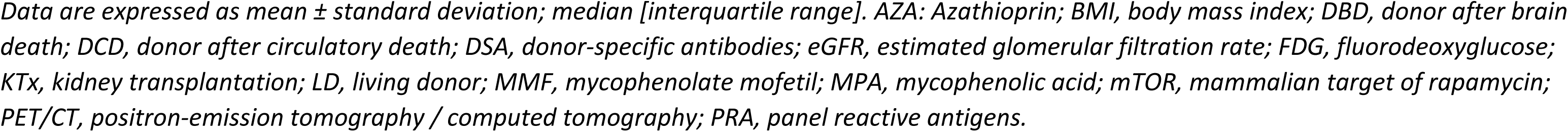
Characteristics of the cohort

#### Histopathology

Biopsies were assessed by pathologists blinded to the results of [18F]FDGPET/CT. Banff 2022 classification was applied. Acute and chronic histological lesions were scored from 0 to 3 on the basis of the severity of infiltration by mononuclear cells in each component (glomeruli (g), peritubular capillaries (ptc), arteries (v), tubules (t) and interstitium (i)). Total inflammation (ti) was scored taking into account interstitial inflammation in both non-sclerotic and sclerotic areas. C4d staining was performed in all cases. The Banff-based categorisation was performed using the raw results of the Banff 2022 classification, which were then integrated into the Banff Automation System for kidney allograft precision diagnostics^17^. This approach allowed us to obtain standardised results.

Biopsies were classified into “normal”; “borderline” (corresponding to Banff category 3: suspicious for acute T-cell-mediated rejection (TCMR), defined by “i=1 and t≥1” or “i=2 and t=1”2, or Banff category 4: C4d staining without evidence of rejection)^2^, or “acute rejection” (corresponding to Banff category 2: active antibody-mediated rejection (AMR), or Banff category 4 : acute TCMR IA or higher). DSA-negative, C4d-negative MVI corresponded to “g+ptc ≥ 2; C4d (-); DSA (-)”^18^. The SCR group was defined as subclinical inflammation including biopsy-confirmed acute rejection and DSA-negative, C4d-negative MVI. BK nephropathies were systematically searched using immunohistochemical detection of SV40 large T antigen in renal allograft biopsies.

#### [^18^F]FDG PET/CT imaging

PET/CT was performed with late acquisitions (180 [180 - 190]minutes) after [^18^F]FDG intravenous injection (222 [193 - 257] Mbq) within a 24-hour period before of after the surveillance biopsy (0.51 [0.47-1.00] day), prior to any modification of immunosuppressive regimens. The mSUV of kidney cortex was measured and averaged from 4 volumes of interest (VOI) distributed in the upper (n=2) and lower (n=2) poles. The mSUV of kidney was normalized to the mSUV of the psoas muscle (VOI of 20 ml) as reference tissue^19^. PET and CT images were acquired using center-specific devices: CHULiège included the cross-calibrated analog GEMINI TF Big Bore and GEMINI TF 16 PET/CT systems (Philips Medical Systems, Cleveland, OH, USA), as well as the digital BIOGRAPH Vision 600 (Siemens); UZA used the digital GE Discovery MI 4R (GE HealthCare). UZB used the cross-calibrated analog BIOGRAPH mCT20 and mCT128 (Siemens).

#### Statistics

The results are presented as means and standard deviations (SD) or medians (Q1 - Q3) for quantitative variables and as frequency tables for qualitative variables. The normality of the parameters was tested using the Shapiro-Wilk test. A logarithmic transformation was applied in cases where the distribution was skewed. To compare parameters between centers, we used the ANOVA test and Scheffé’s multiple comparisons (means), the Kruskal-Wallis test and Dwass, Steel, Critchlow-Fligner (DSCF) multiple comparisons (medians), and the chi-square test and Bonferroni correction for multiple comparisons (proportions). The correlation between the mSUVR and Banff “ti” score was assessed using 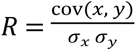. To study the risk of biopsy-proven AR (including AMR, TCMR, and MVI) based on mSUVR, a stratified logistic regression by center was used and odds ratios (OR) and 95% confidence intervals (95%CI) were reported. Stratified logistic regression models by center were then performed by forcing the SUV value or ratio and using stepwise selection of parameters with a p-value < 0.10 in univariate analysis. The sample size calculation on the basis of a 7% incidence of SCR^16^, with α/β errors of 5% and 80%, respectively, estimated that 156 [^18^F]FDG PET/CTwere needed to statistically test its diagnostic performance in biopsy-proven SCR detection. Results were considered significant at a 5% uncertainty level (p<0.05). Calculations were performed using SAS version 9.4.

## Results

From January 2021 to March 2025, we prospectively performed 191 [^18^F]FDG PET/CTin adult KTRs who underwent surveillance transplant biopsy at 91 [87- 98] days *post* KTx. Six cases were excluded from the stats because of BK polyomavirus viremia > 3 log_10_ copies (n=4) or uninterpretable histology (n=2) (**Figure 1**). The mean age of the 185-KTR cohort was 55.0±12.8 years with a sex ratio (M/F) of 1.76 and a mean body mass index (BMI) of 25.7±4.4 kg/m^2^. The mean age of donors was 48.4±12.3 years with a sex ratio of 1.53 and a mean BMI of 25.8±4.6 kg/m^2^ (**Table 1**). Centers statistically differed by (i) the cold ischemia time (421 [258 - 747] minutes, p=0.0005); (ii) the median interval between KTx and transplant biopsy (91 [87–98] days, p=0.0004); serum creatinine levels (1.31 [1.07–1.54] mg/dL, p=0.013); and urinary protein/creatinine ratio (119 [81–190] mg/g, p=0.0001) (**Supplementary Table 1**).

**Figure 1:**
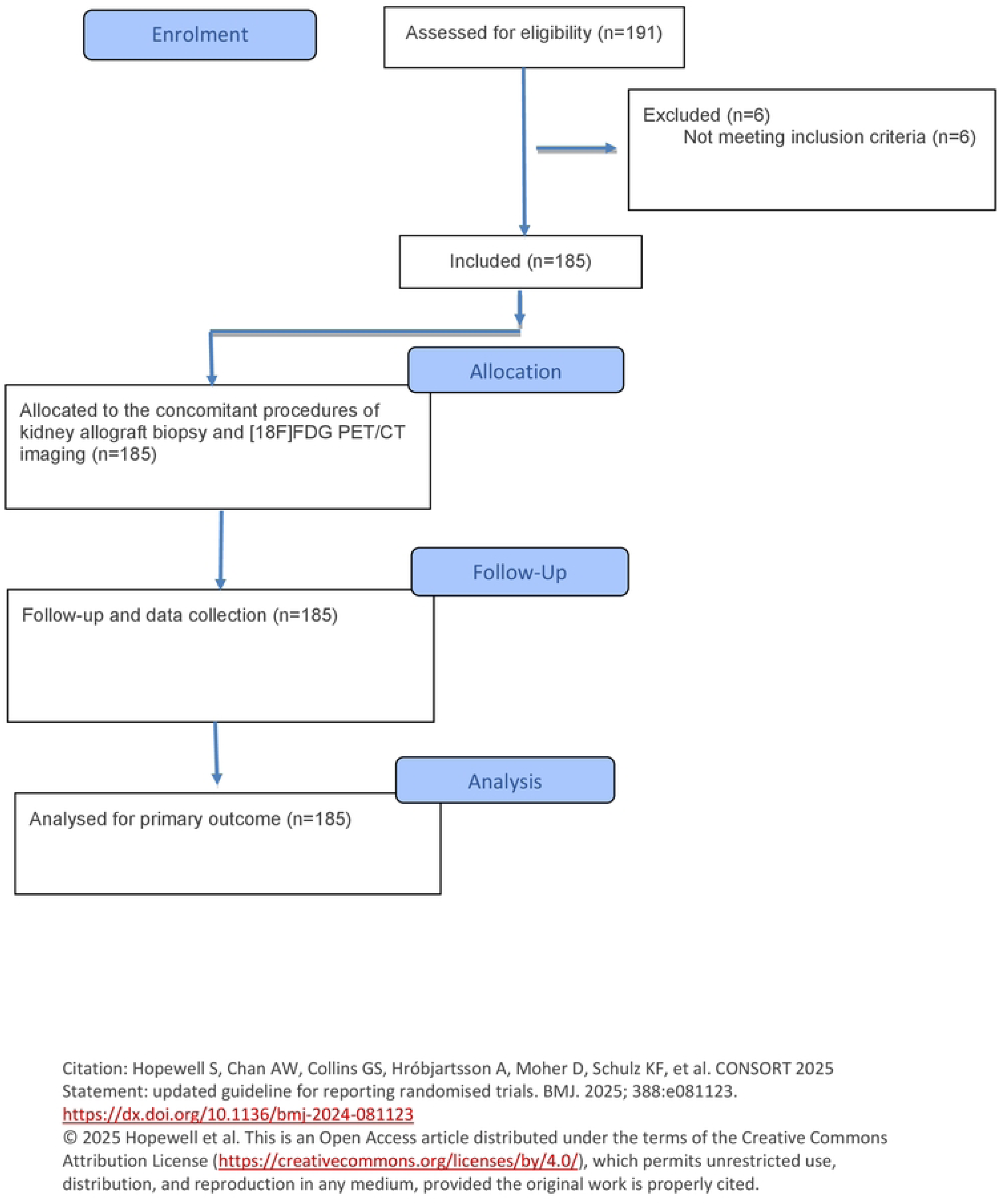
CONSORT 2025 Flow Diagram Between January 2021 and March 2025, 191[^18^F]FDG PET/CT were prospectively performed at the time of surveillance transplant biopsy at ∼3 months *post* transplantation. Six cases were excluded: 4 PCR-proven BK nephropathies and 2 uninterpretable histology.

The 185-KTR cohort was categorized upon Banff 2022-based histology: normal (n=158); borderline (n=18); SCR (n=9, including 6 T-cell-mediated rejection (TCMR-1A) and 3 DSA-negative C4d-negative MVI) (**Supplementary Figure 1**). Immunoreactivity against C4d was detected in 1 case with no features of AMR and no DSA (i.e. “isolated C4d positivity”). No AMR was diagnosed in our surveillance cohort. No significant correlation was observed between the mSUVR and ti score (R=0.032, p-value=0.67) or acute composite Banff score (i.e. the sum of g, ptc, t, i, and v; R²=0.002, p-value=0.54). The mSUVR reached 2.33 [1.97 - 2.93], 2.71 [2.50 -3.33] and 2.42 [2.27 - 3.14] in normal, borderline and SCR groups, respectively (**Figure 2**). In univariate analysis stratified by center, the risk of non-normal histology (n=27, including Borderline and SCR) increased with donor age (OR=1.06 [1.01 - 1.10], p=0.009), serum creatinine level (OR=6.15 [1.17 - 32.42], p=0.032), and mSUVR (OR=5.16 [1.21 - 22.11], p=0.017) (**Supplementary Table 2**). In multivariate models stratified by center, mSUVR was not significantly associated with the risk of non-normal histology (OR=4.11 [0.91–18.49, p=0.066). Furthermore, the risk of biopsy-proven SCR (n=9) was not significantly associated with any clinical, biological or radiological parameter, including mSUVR, in univariate models stratified by center. The individual mSUVR of the TCMR subgroup were: 1.77, 2.12, 2.27, 2.28, 2.14, and 3.79. The mSUVR of the MVI subgroup were: 2.42, 2.74, and 3.99. Focusing on the previously proposed >2.4 mSUVR threshold^16^, no significant difference was observed between the proportion of biopsy-proven SCR above the threshold (5/9, 55.6%) and the proportion of normal or borderline histologies above the threshold (87/176, 49.4%) (p=0.75).

**Figure 2:**
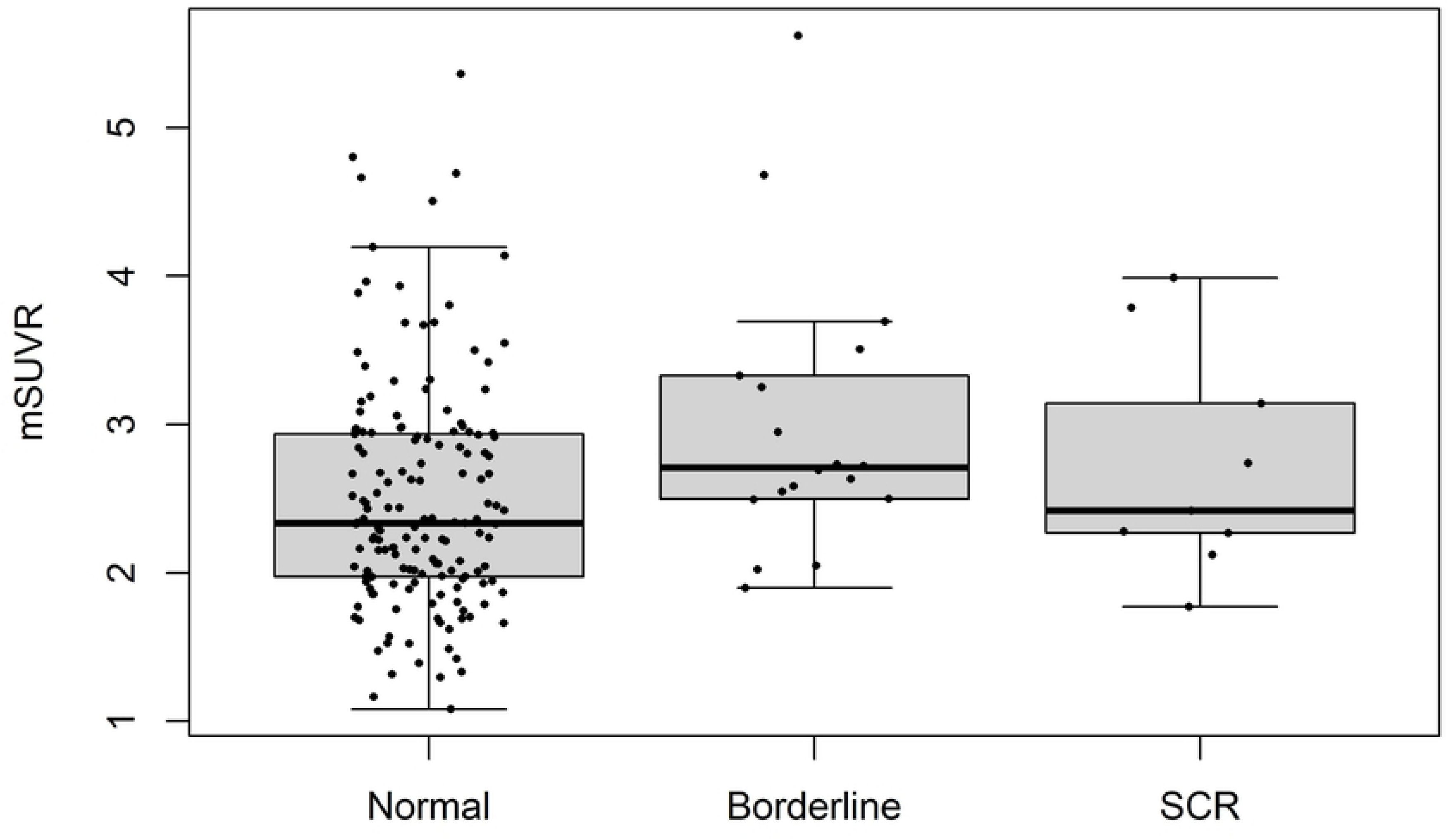
The kidney/psoas ratio of SUV_mean_ (mSUVR) upon histopathological categories The 185-KTR cohort was categorized upon Banff 2022-based histology: normal (n=158); borderline (n=18); subclinical rejection (SCR, n=9). The mSUVR reached 2.33 [1.97 - 2.93], 2.71 [2.50 -3.33] and 2.42 [2.27 - 3.14] in normal, borderline and SCR groups, respectively

## Discussion

The term “SCR” was introduced by Rush et al. in the 1990’s to refer to stable allografts displaying an interstitial infiltrate and tubulitis^20^. The diagnosis of SCR is clinically relevant, given the efficiency of early treatment of SCR to decrease the incidence of late allograft rejection episodes and the occurrence of fibrosis, thereby improving long-term graft function in KTR^3^. However, the systematic indication of surveillance biopsy remains debatable, and it is not currently part of the KDIGO 2009 guidelines, rather emphasizing broad justifications for per-cause biopsy^21^. The development and validation of alternative non-invasive approaches “ruling out” SCR may help avoid biopsy-associated complications and limitations in stable patients with a low risk of SCR^22^. In a pilot prospective monocentric 92-KTR cohort, we postulated that a ratio of kidney/psoas mSUVR lower than 2.4 would non-invasively exclude SCR, with a negative predictive value of 98%^16^. In the present multicenter validation study, we do not confirm that mSUVR reliably exclude SCR in stable non-sensitized KTR at 3 months *post* KTx. Indeed, the median mSUVR in the group with biopsy-proven SCR, including 6 TCMR and 3 MVI, reached 2.42 [2.27 - 3.14]. The subcategory of "MVI, DSA-negative, C4d-negative” was introduced in the Banff 2022 because of its significant risk of graft failure^23^. Typically, MVI is an histological feature of immune-mediated injury at the capillary interface of renal allografts, characterized by immune cell infiltration into glomerular and peritubular capillaries. Emerging evidence from transcriptome analyses highlights natural killer cells as possible effectors, regardless of DSA status. No AMR was diagnosed in the present series. The “Borderline” category is a well-known limitation of the Banff classification, with recent controversy about the tubulitis without interstitial inflammation (i.e. “i0 tx”)^24^. In 2009, the “Borderline” definition included foci of tubulitis (t1, t2, or t3) with minor interstitial infiltration (i0 or i1) or interstitial infiltration (i2, i3) with mild (t1) tubulitis. By contrast, the Banff 2019 classification narrowed this definition as follows: tubulitis (t1, 2, or 3) and interstitial inflammation (i1) or tubulitis (t1) and interstitial inflammation (i2 or i3) (https://banfffoundation.org). Molecular microscopy and omics are currently ongoing to better specify this “Borderline” phenotype from both diagnostic and prognostic perspectives^25^. Similarly, one may hypothesize that non-invasive biomarkers and/or imaging-based markers may help better distinguish “borderline/normal” lesions from “borderline/rejection” lesions by reflecting the *in toto* intensity of the inflammation within the kidney allograft. In our cohort, the mSUVR of kidney allograft with “borderline histology” reached 2.71 [2.50 -3.33], which was significantly higher than the “normal histology” (2.33 [1.97 - 2.93], p=0.022). Recent investigations have highlighted the continuous nature of the rejection process regardless of the underlying disease cause, which questions the dichotomization of the Banff classification^26^.

The limitations of our study include the low incidence of pathological cases, although one may argue that a 13% incidence of “Borderline + SCR” cases is consistent with 10-20% range reported from most studies under modern immunosuppression in the early post-transplant course. Still, we have to admit that, despite the small number of cases, [18F]FDGPET/CT cannot reliably rule out SCR, since 5/6 TCMR showed mSUVR below the 2.4 threshold established by our previous study^16^. Given the multicentric design of the present trial, the quantification of [^18^F]FDG uptake in the renal allograft was prospectively performed in each center according to the above-detailed protocol. The lack of centralized review of [^18^F]FDG PET/CT images is a potential limitation of our pragmatic design. Still, the use of multiple independent 1-ml VOI distributed in lower and upper renal poles aimed to avert sampling errors and consider a large representative zone of renal parenchyma. The use of the mSUVR referenced to the psoas muscle aimed to further reduce the variability of the radiotracer biodistribution between (i) patients and (ii) PET/CT devices^19,27^. Exams were indeed performed on both analog and digital [^18^F]FDG PET/CT systems^28^, the latter, based on silicon photomultipliers, providing superior resolution, sensitivity, and improved image quality. The stats were therefore stratified by center. The nature of the radiotracer [^18^F]FDG explains *per se* the poor specificity of [^18^F]FDGPETCT in the differential diagnosis of inflammatory conditions. Radiomics is the high-throughput extraction and analysis of quantitative features from medical images to characterize tissue phenotypes. Radiomics has been increasingly used in inflammatory and infectious diseases, such as sarcoidosis and vasculitis^29,30^. One may hypothesize that radiomics, including (semi-) automatic segmentation methods of the renal cortex, may improve the diagnostic yield of [^18^F]FDG PET/CT in SCR screening. Combining high negative predictive value with high sensitivity would ensure a low false negative rate when considering both the population- and disease-specific performance, and make a more convincing case for avoidance of unnecessary biopsies.

In conclusion, our prospective multicentric cohort including 185 cases combining transplant biopsy and [^18^F]FDG PET/CT at 3 months *post* kidney transplantation did not validate the 2.4 mSUVR threshold to safely exclude SCR. Further investigations are warranted to test the utility of innovative imaging-based markers in the follow-up of KTR.

## Data availability statement

The data underlying this study are available from the corresponding author upon reasonable request. Due to institutional regulations and the protection of participant privacy, individual-level clinical data cannot be made publicly available but may be shared in anonymized form for research purposes subject to appropriate ethical approvals.

## Data Availability

All relevant data are within the manuscript and its Supporting Information files.

## Acknowledgements

The authors cordially thank the patients-participants, the surgeons (A. De Roover, O. Detry, N. Gilbo, N. Meurisse, and M. Vandermeulen), the physicians (L. Vanovermeire and P. Xhignesse), and the members of the local transplant coordination center (J Mornard, F. Sinte and A Warmoes) for their commitment to kidney transplantation at the University of Liege Hospital in Liege, Belgium; the surgeons (T. Chapelle, G. Roeyen, B. Bracke, V. Hartman, B. Hendrikx, E. Liekens), the physicians (M. Dirix, H. de Fijter, V. de Meier), the study coordinator (P. Lievens) and transplant coordinators (G. Van Beeumen, P. Hollants, A. Van de Weyer, L. Van den Bergh, M. De Deyne) at Universitair Ziekenhuis Antwerpen in Antwerp, Belgium; the surgeons, the physicians, the study coordinator and the transplant coordinators at Universitair Ziekenhuis Brussel in Brussels, Belgium. AB is a Fellow of the Fonds National de la Recherche Scientifique. FJ and RH received support from the University of Liège (Fonds Spéciaux à la Recherche; Fondation Léon Fredericq), as well as from ULiège Academic Hospital (Fonds d’Investissement pour la Recherche Scientifique, FIRS). This study was supported by the Belgian Transplant Society.

## Authors‘ contributions

- PL, AB, AM, LP, LW, RH and FJ participated in research design
- PL, AB, AM, LP, RH and FJ participated in the writing of the paper
- PL, AB, AM, LP, CB, LC, HE, AJ, AD, CG, SG, PE, RH, LW, RH and FJ participated in the performance of the research
- PL, AB, AM, LP, LS and FJ participated in data analysis

